# Identification of a protein expression signature distinguishing early from organising diffuse alveolar damage in COVID-19 patients

**DOI:** 10.1101/2022.12.09.22283280

**Authors:** Helen Ashwin, Luke Milross, Julie Wilson, Joaquim Majo, Jimmy Tsz Hang Lee, Grant Calder, Bethany Hunter, Sally James, Dimitris Lagos, Nathalie Signoret, Andrew Filby, Omer Ali Bayraktar, Andrew J. Fisher, Paul M. Kaye

## Abstract

Diffuse alveolar damage (DAD) is a histopathological finding associated with severe viral infections, including SARS-CoV-2. However, the mechanisms mediating progression of DAD are poorly understood. Applying protein digital spatial profiling to lung tissue obtained from a cohort of 27 COVID-19 autopsy cases from the UK, we identified a protein signature (ARG1, CD127, GZMB, IDO1, Ki67, phospho-PRAS40 (T246), and VISTA that distinguishes early / exudative DAD from late / organising DAD with good predictive accuracy. These proteins warrant further investigation as potential immunotherapeutic targets to modulate DAD progression and improve patient outcome.

## INTRODUCTION

The COVID-19 pandemic has claimed over 6.6 million lives and despite vaccines that prevent serious illness and use of dexamethasone in severely ill patients, worldwide deaths continue to accrue^1^. There is, therefore, a continued need to identify new treatment options to minimise disease severity. Diffuse alveolar damage (DAD) reflects a continuum of immunopathology associated with multiple causes of lung injury and is a primary histological feature of fatal COVID-19^2, 3^. However, the cellular and molecular pathways associated with the progression of DAD from its early exudative phase (EDAD), characterised by oedema, hyaline membranes, and inflammation, to a late organising and loosely fibrotic phase (ODAD) remain unclear.

To begin to address this question, we examined lung tissue from a cohort of COVID-19 autopsy cases in the UK. We used digital spatial profiling (DSP) to determine differences in protein expression between regions of interest identified histologically as EDAD or ODAD. We focused on protein targets with therapeutic potential demonstrated in other diseases and/or pre-clinical models to identify potential candidates for re-purposing in COVID-19.

## MATERIALS AND METHODS

Lung tissue from patients that had died with SARS-CoV-2 was selected from a larger cohort assembled by the UK Coronavirus Immunology Consortium (UK-CIC). A full description of the UK-CIC cohorts will be provided elsewhere (Milross et al, ms in preparation). Patients selected for the current study (5 female, 22 male; 7 black/Asian/minority ethnic, 20 caucasian) had histological evidence of DAD (online supplemental table 1) without concurrent bronchopneumonia or histology attributable to acute cardiac failure. Regions (approx. 600μm^2^) reflecting EDAD, ODAD or a mixed phenotype (MDAD) were identified by a pathologist with cardiothoracic expertise on H&E-stained FFPE sections and used to guide subsequent ROI selection for GeoMx^®^ DSP. Digital counts reflecting protein expression were analysed using unsupervised methods for dimensionality reduction, methods for class discrimination and using linear mixed modelling (see online supplementary methods).

## RESULTS

We examined 194 ROIs (7 ± 2 ROIs per patient; 122 EDAD, 50 ODAD, 22 MDAD; **figure 1A** and online supplemental table 1). Principal components analysis (PCA) showed separation of each form of DAD with 41.4% of variance accounted for by PC1 and PC2 (**figure 1B and C**). We next applied partial least squares regression (PLS-R) (**figure 2A**) and identified variables responsible for group separation using variable importance in projection (VIP) scores. Proteins with VIP scores > 1.3 (ARG1, CD127, CD163, GZMB, IDO1, Ki67, phopsho-PRAS40 (T246) and VISTA; **figure 2B**) largely mirrored what was observed with PCA. These 8 variables were used to classify ROIs in PLS linear discriminate analysis (PLS-LDA) with leave-one-patient-out (LOPO) cross validation to prevent overfitting. This achieved a predictive accuracy of 93% and 80% for EDAD and ODAD respectively (**figure 2C**). MDAD ROIs were consistently mis-classified, likely a reflection of heterogeneity and the transitional nature of the pathology within this group. Finally, we generated a Receiver Operating Characteristic (ROC) curve for EDAD and ODAD samples using LOPO-cross validation in PLS-R, showing the predictive accuracy as the discriminatory threshold is varied (f**igure 2D**).

**Figure 1.**
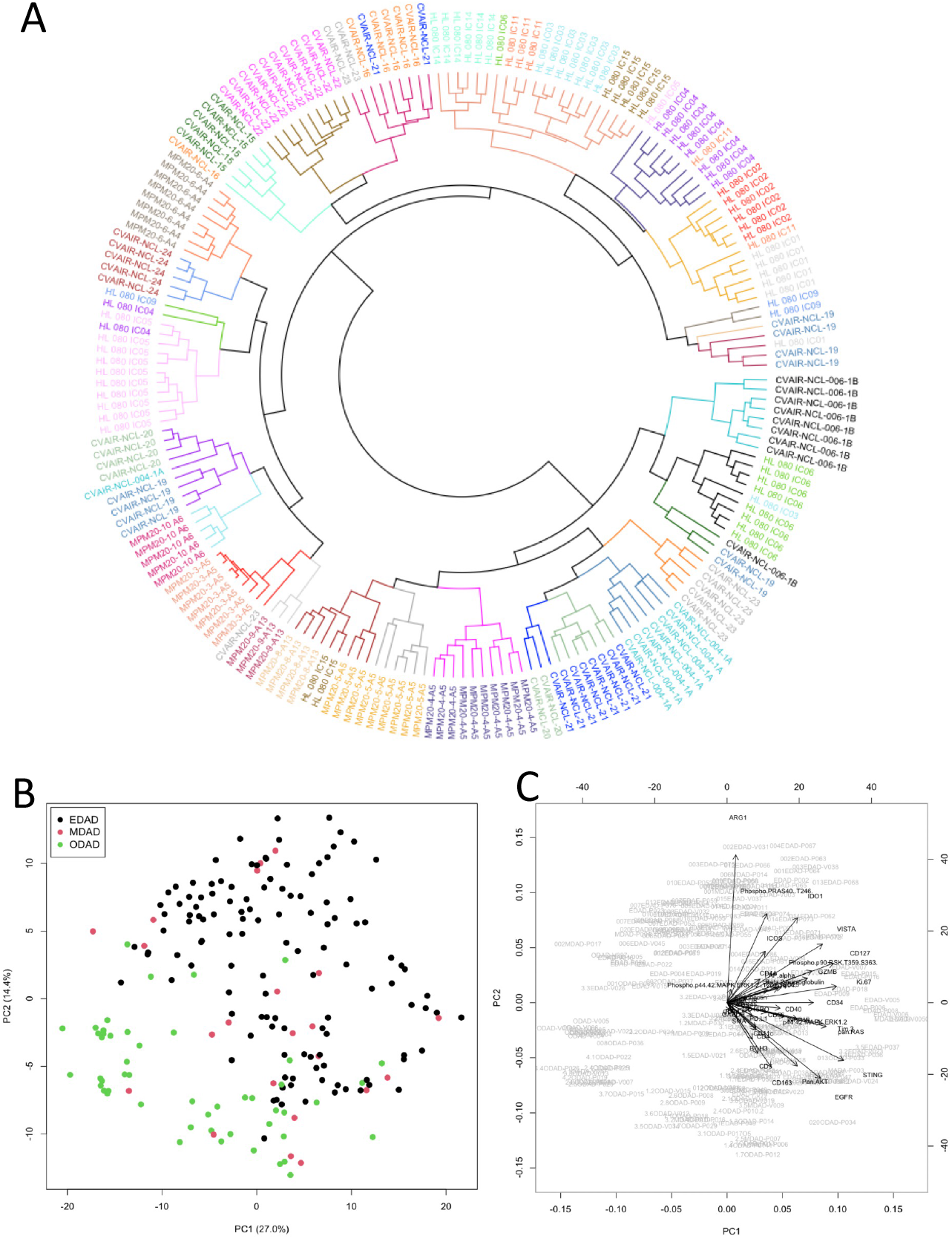
Analysis of protein DSP data. **A**, Circular dendrogram from hierarchical clustering of protein DSP ROIs (with patient identifiers colour-coded and clusters coloured separately; see supplementary table 1). **B and C**, Principal Component Analysis (PCA) scores plot for the first two principal components coloured by EDAD, MDAD and ODAD (B) and with loadings shown as vectors (C).

**Figure 2.**
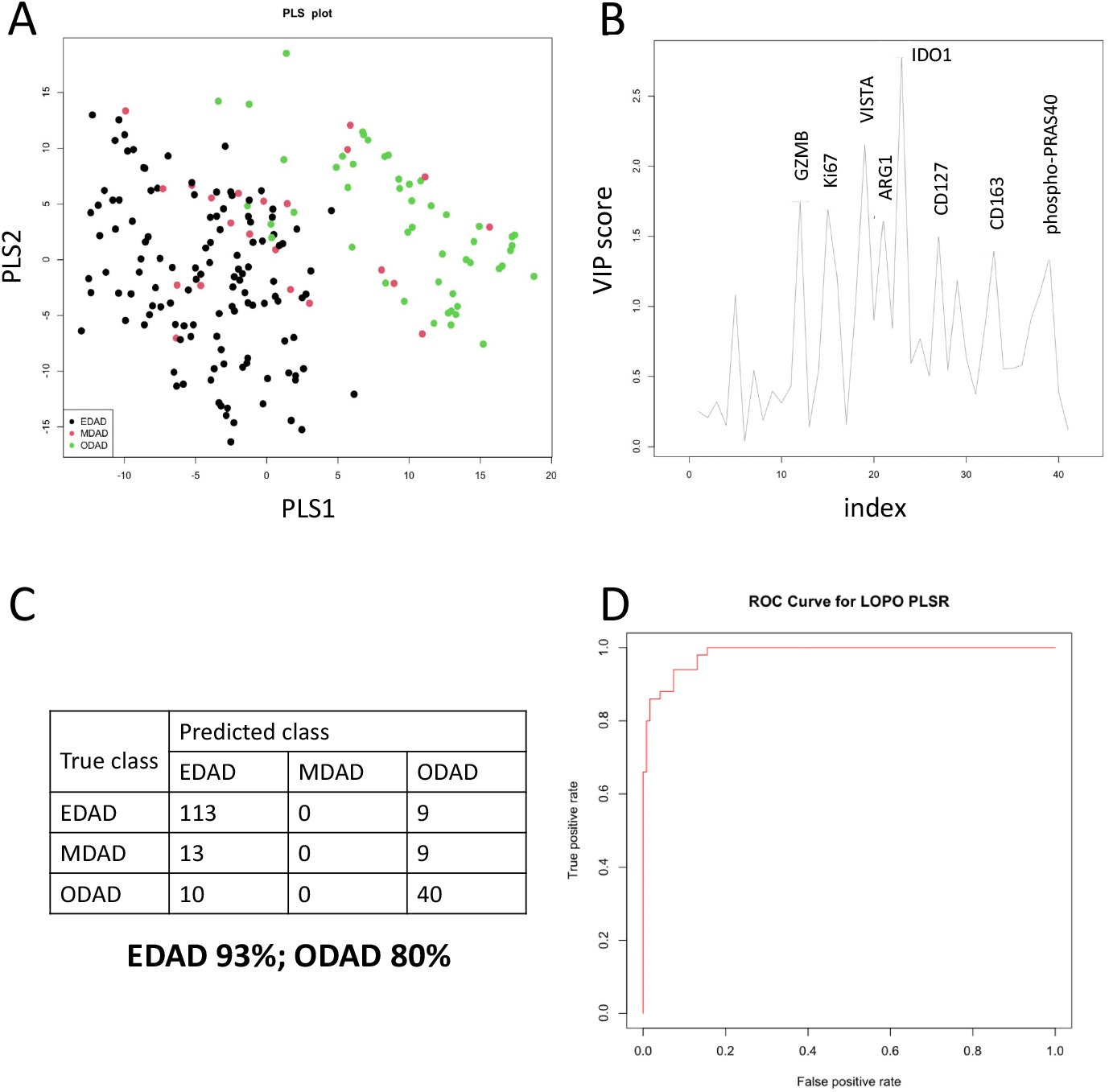
Discrimination of DAD classes based on protein signature. **A and B**, Partial Least Squares Analysis of EDAD, MDAD and ODAD samples shown as PLS plot (A) and by Variable Importance in Projection (VIP) score (B). **C**, Confusion matrix for results of PLS-LDA leave one patient out prediction using 8 variables with VIP scores > 1.3 (GZMB, Ki.67, VISTA, ARG1, IDO1, CD127, CD163, Phospho.PRAS40). **D**, Receiver Operating Characteristic (ROC) curve generated for EDAD vs ODAD ROIs.

We independently analysed these data using linear mixed modelling to account for potentially confounding factors (including repeat measures and cohort effects) and identified eleven targets (ARG1, B2M, CD14, CD34, CD44, CD127, GZMB, IDO1, Ki67, phospho-PRAS40 (T246) and VISTA) distinguishing EDAD and ODAD (>1.5-fold change cut off, FDR = 5%; **figure 3A, C**). MDAD was similarly distinguished from ODAD (**figure 3B, C**), but no proteins were significantly different between EDAD and MDAD.

**Figure 3.**
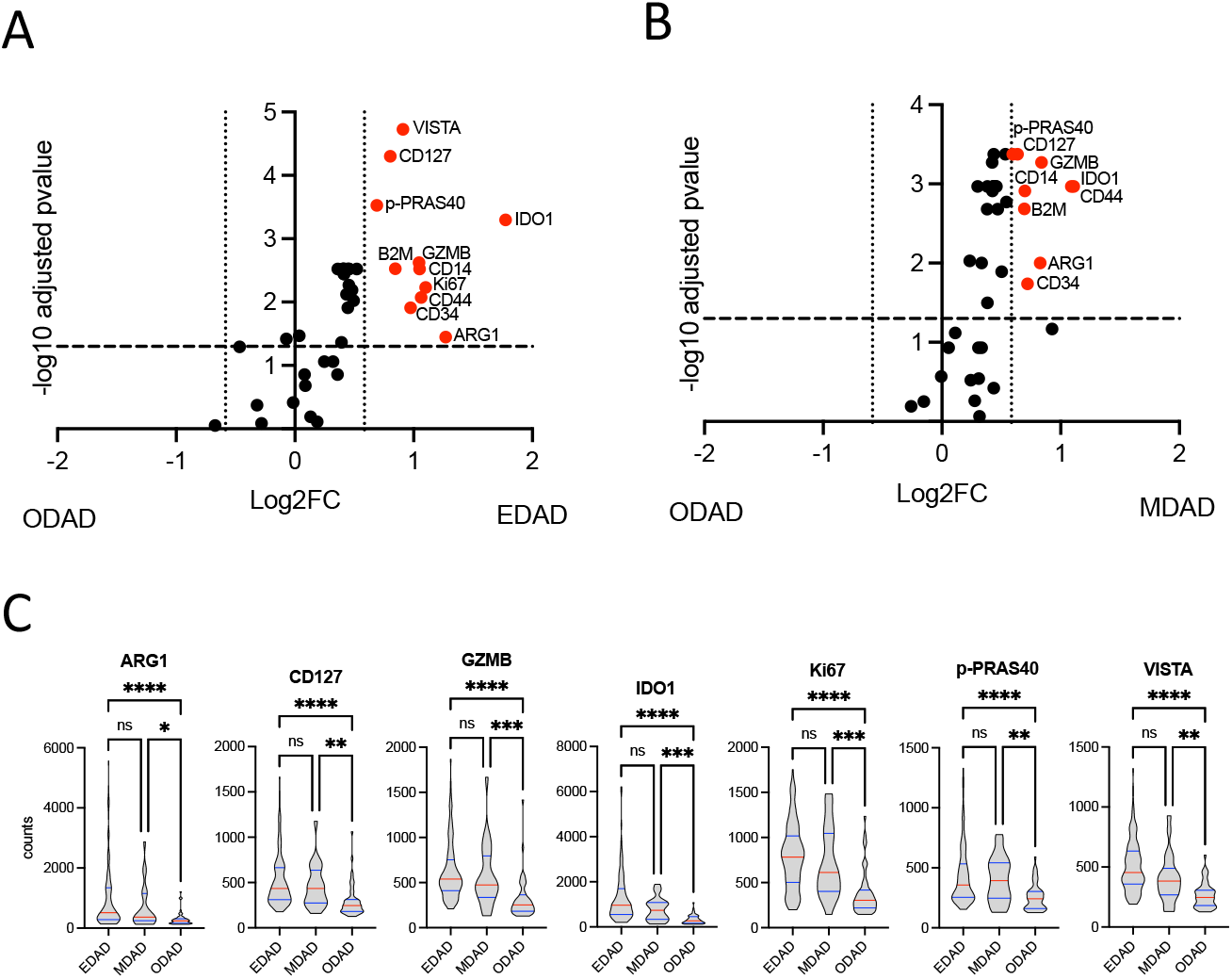
Differential target expression between EDAD and ODAD using linear mixed modelling. **A and B**, Differentially expressed (FDR 5%; FC = 1.5) protein targets between EDAD and ODAD (A) and MDAD and ODAD (B). Data derives from a Linear mixed modelling with patient repeat measures and cohort as a random effect. **C**, Individual ROI counts for EDAD, MDAD and ODAD ROIs for identified target proteins. ns, non-significant; **, p<0.01; ***, p<0.001; **** p<0.0001 between indicated groups.

Collectively, our data suggest a core protein signature comprising ARG1, CD127, GZMB, IDO1, Ki67, phospho-PRAS40 (T246), and VISTA distinguishes EDAD from ODAD ROIs in this patient group (**figure 2C**). Nevertheless, our data also suggest further patient heterogeneity within EDAD ROIs. This was most marked for ARG1, which was absent from all EDAD ROIs in 8/20 patients. Although sample size precluded a formal analysis, this appeared unrelated to sex, place of death, duration of disease or cohort (online supplementary table 1).

## DISCUSSION

Using DSP to interrogate well-annotated lung tissue, we identified a core protein signature discriminating early from late phases of DAD. Not surprisingly given the targeted nature of our panel, the proteins we identified have well known functions in inflammation and immunity, but they have not previously been evaluated in relation to DAD progression.

ARG1 is elevated in the lungs of severe COVID-19 patients, being expressed by CD11b^+^CD66b^+^ granulocytic myeloid-derived suppressor cells.^4^ IDO1 was detected in lung tissue in another autopsy series, though most tryptophan-catabolizing activity was associated with IDO2.^5^ CD127 expression on monocytes has been noted at sites of hyperinflammation^6^, whereas VISTA has been proposed as a therapeutic target to minimise inflammation.^7^ Finally, phosphorylation of PRAS40 at T246 releases mTORC1 to perform its many downstream functions and elevated phopsho-PRAS40 (T246) has been used as a biomarker of PI3K/Akt/mTORC1 activation^8^, a pathway implicated in idiopathic pulmonary fibrosis and DAD^9^.

This study has limitations: 1. DSP quantifies protein expression across the entire ROI and cannot distinguish multiple cells with low target expression vs. few cells with high expression; 2. our patient cohort was too small to perform sub-group analysis based on age, gender, disease duration, or place of death; 3. we cannot rule out that patients had other forms of concurrent disease or different forms of DAD in other areas of lung not sampled here and this may account for some of the inter-patient heterogeneity observed; 4. further validation is required in an independent patient cohort, preferably incorporating single cell technologies.

Notwithstanding these limitations, to our knowledge this is the first study to apply highly multiplexed DSP to discriminate between EDAD and ODAD. The extent to which the many millions of COVID-19 survivors are at risk of developing pulmonary fibrosis is only beginning to be understood^10^. Importantly, many of the protein targets we have identified as being highly expressed at the early stages of DAD are amenable to therapeutic intervention with existing drugs or drugs in development. Hence further exploration of these targets in pre-clinical models of SARS-CoV-2 infection could provide an evidence base on which to base future intervention trials.

## Data Availability

Raw DSP count data are deposited at the Open Science Framework and are available at https://osf.io/69znm/

## Acknowledgements

The authors would like to acknowledge the tissue donors and their families for their contribution to medical science.

## Contributors

AJF, AF, OAB, DL, JTHL, NS and PMK contributed to the conception and design of the study. HA, GC, AF, BS, and SJ generated samples and performed DSP analysis; LM and JM performed pathology guided ROI selection; JW and PMK conducted data analysis. All authors contributed to drafting the manuscript or critical review and all provided approval for submission.

## Ethical Approval

Human samples used in this research project were partly obtained from the Newcastle Hospitals CEPA Biobank and their use in research is covered by Newcastle Hospitals CEPA Biobank ethics – REC 17/NE/0070. Additional human samples used in this research project were obtained from the Imperial College Healthcare Tissue Bank (ICHTB). ICHTB is supported by the National Institute for Health Research (NIHR) Biomedical Research Centre based at Imperial College Healthcare NHS Trust and Imperial College London. The views expressed are those of the author(s) and not necessarily those of the NHS, the NIHR or the Department of Health. ICHTB is approved by Wales REC3 to release human material for research (22/WA/0214). Additional human samples used in this research project were obtained from the ICECAP tissue bank of the University of Edinburgh. ICECAP is approved by the East of Scotland Research Ethics Service to release human material for research (16/ED/0084). Analysis of these samples at the University of York was approved by the Hull York Medical School Ethics Committee (20/52).

## SUPPLEMENTAL MATERIALS

**Online supplementary table 1** – Patient demographics and DSP counts data file (Excel)

**Online supplementary methods**

## ONLINE SUPPLEMENTARY METHODS

### Patient samples

Autopsy lung tissues from 27 patients were obtained from biobanks at the University of Newcastle, University of Edinburgh, and Imperial College London. Patients were from both the first and second wave of the UK pandemic. These patients were selected from a larger histopathological study on the basis that they showed DAD in the absence of additional lung complications associated with pneumonia or heart failure. A description of the full cohort (demographics, time to death, comorbidities etc) will be published elsewhere (Milross et al, ms in preparation).

### Nanostring GeoMx protein spatial profiling

4μm thick FFPE lung sections were used for protein spatial profiling using the Nanostring GeoMx^®^ platform. Slides were stained with CD3 and CD68 as morphological markers and with a panel of 68 oligo-nucleotide conjugated antibodies comprising the Immune Cell Profiling Core (24 Abs), IO Drug Target Panel (10 Abs), Immune Activation Status Panel (8 Abs), Immune cell Typing Panel (7 Abs), PI3K/AKT Signalling Panel (9 Abs) and the MAPK Signalling Panel (10 Abs). Region of interest (ROI) selection was pathologist-guided, based on the examination of H&E-stained serial sections. Regions conforming the histological description of exudative and organising DAD were identified in each patient’s lung tissue. ROI capture was performed using a GeoMx Spatial profiler instrument (Nanostring, Seattle, WA, USA).

Digital count data were normalised to positive ERCC controls and to housekeeping controls (GAPDH and Histone H3). Housekeeping targets were selected based on high correlation with isotype controls. ROIs with abnormal levels of hybridisation, HK expression or low isotype control background were removed from the analysis. Targets were removed from analysis if signals were below the geometric mean of the isotype controls. Data was exported for further analysis in R (see below) and analysed using linear mixed modelling using GeoMx software (version 2.0) with patient ID and cohort selected as random variables. Volcano plots were generated in GeoMx^®^ software and show significance scores with FDR correction (5%) based on Benjamini, Krieger, and Yekutieli two stage set-up method and Log2 fold change cut-off of 0.589 (1.5-fold change).

### Statistical analysis

Statistical analyses were carried out in R version 4.1.1.^1^ The base R function prcomp was used for PCA, while the pls package ^2^ was used for PLSR. Classification was performed using the plsgenomics R package ^3^ with LOPO cross-validation to avoid overfitting in this supervised approach. Here all ROIs for each patient in turn were left out and the remaining data used to build the model which was then used to predict the class of the left-out ROIs. Results are shown for the ROIs that were not used in model training. In order to show the predictive accuracy as the discriminatory threshold was varied, a receiver operator curve (ROC) was generated using the R package ROCR ^4^.

## REFERENCES

1. Centre, J.H.C.R. [cited 23/11/22] Available from: https://coronavirus.jhu.edu/map.html

2. Li, Y. et al. Progression to fibrosing diffuse alveolar damage in a series of 30 minimally invasive autopsies with COVID-19 pneumonia in Wuhan, China. Histopathology 78, 542–555 (2021).

3. Milross, L. et al. Post-mortem lung tissue: the fossil record of the pathophysiology and immunopathology of severe COVID-19. Lancet Respir Med 10, 95–106 (2022).

4. Dean, M.J. et al. Severe COVID-19 Is Characterized by an Impaired Type I Interferon Response and Elevated Levels of Arginase Producing Granulocytic Myeloid Derived Suppressor Cells. Front Immunol 12, 695972 (2021).

5. Guo, L. et al. Indoleamine 2,3-dioxygenase (IDO)-1 and IDO-2 activity and severe course of COVID-19. J Pathol 256, 256–261 (2022).

6. Zhang, B. et al. CD127 imprints functional heterogeneity to diversify monocyte responses in inflammatory diseases. J Exp Med 219 (2022).

7. ElTanbouly, M.A. et al. VISTA: A Target to Manage the Innate Cytokine Storm. Front Immunol 11, 595950 (2020).

8. Laplante, M. & Sabatini, D.M. mTOR signaling in growth control and disease. Cell 149, 274–293 (2012).

9. Saito, R. et al. Immunohistochemical evidence for the association between attenuated mTOR signaling and diffuse alveolar damage, a fatal lung complication. Tohoku J Exp Med 234, 67–75 (2014).

10. Tarraso, J. et al. Lung function and radiological findings 1 year after COVID-19: a prospective follow-up. Respir Res 23, 242 (2022).

## Supplementary references

1. R Core Team (2021). R: A language and environment for statistical computing. R Foundation for Statistical Computing, Vienna, Austria. URL https://www.R-project.org/.

2. Kristian Hovde Liland, Bjørn-Helge Mevik and Ron Wehrens (2021). pls: Partial Least Squares and Principal Component Regression. R package version 2.8-0. https://CRAN.R-project.org/package=pls

3. Anne-Laure Boulesteix, Ghislain Durif, Sophie Lambert-Lacroix, Julie Peyre and Korbinian Strimmer. (2018). plsgenomics: PLS Analyses for Genomics. R package version 1.5-2. https://CRAN.R-project.org/package=plsgenomics.

4. Sing T, Sander O, Beerenwinkel N, Lengauer T (2005). ROCR: visualizing classifier performance in R. Bioinformatics, 21(20), pp.3940–3941. https://CRAN.R-project.org/package=ROCR

